# Functional connectivity directionality between large-scale resting-state networks in children and adolescence from the Healthy Brain Network sample

**DOI:** 10.1101/2020.10.09.20207936

**Authors:** Martina J. Lund, Dag Alnæs, Jaroslav Rokicki, Simon Schwab, Ole A. Andreassen, Lars T. Westlye, Tobias Kaufmann

## Abstract

Mental disorders often emerge during adolescence and have been associated with age-related differences in connection strengths of brain networks (static functional connectivity), manifesting in non-typical trajectories of brain development. However, little is known about the direction of information flow (directed functional connectivity) in this period of functional brain progression. We employed dynamic graphical models (DGM) to estimate directed functional connectivity from resting state functional magnetic resonance imaging data on 1143 participants, aged 6 to 17 years from the healthy brain network (HBN) sample. We tested for effects of age, sex, cognitive abilities and psychopathology on estimates of direction flow. Across participants, we show a pattern of reciprocal information flow between visual-medial and visual-lateral connections, in line with findings in adults. Investigating directed connectivity patterns between networks, we observed a positive association for age and direction flow from the cerebellar to the auditory network, and for the auditory to the sensorimotor network. Further, higher cognitive abilities were linked to lower information flow from the visual occipital to the default mode network. Additionally, examining the degree networks overall send and receive information to each other, we identified age-related effects implicating the right frontoparietal and sensorimotor network. However, we did not find any associations with psychopathology. Our results revealed that the directed functional connectivity of large-scale brain networks is sensitive to age and cognition during adolescence, warranting further studies that may explore trajectories of development in more fine-grained network parcellations and in different populations.

## Introduction

The brain undergoes tremendous changes throughout life, where childhood and adolescence is a particularly sensitive developmental period for brain maturation processes (Blakemore, 2012). A vital part of the brain’s maturation happens in the functional networks that show pronounced reorganization in order to facilitate neural efficiency and integration of information, as indicated by functional magnetic resonance imaging (fMRI) studies in healthy children and young individuals as well as in clinical populations in the same age range (Insel, 2010; Keshavan, Giedd, Lau, Lewis, & Paus, 2014; Kolskar et al., 2018; Rausch et al., 2016; Schweinsburg, Nagel, & Tapert, 2005). This supports that this time period is a sensitive phase, where there is large alterations in the functional interconnections between brain regions, denoting the functional connectome, that link to when different mental disorders emerge (Insel, 2010).

The primary method to study the functional connectome so far has been to estimate static functional connectivity between brain regions, where research has shown that the functional interconnections in healthy children and adolescents become more specialized (Li, Deng, He, Zhai, & Jia, 2019). Resting-state networks (RSNs) also become more coherent and stable during this time-period (Hoff, Van den Heuvel, Benders, Kersbergen, & De Vries, 2013), and evidence has indicated aberrant connectivity in individuals with pre-clinical psychiatric symptoms (Kaufmann et al., 2017), which is in line with the brain dysconnectivity hypothesis in mental disorders (Connolly et al., 2013; Di Martino et al., 2011; Friston & Frith, 1995; Hamm et al., 2014). Recently, estimating connectivity direction (Friston, Moran, & Seth, 2013) has received interest as a field, as it could give new knowledge about the connections between brain regions by yielding insight into the directionality in neural information flow. For instance, Riley et al. (2018) examined connectivity direction with the use of task-fMRI in a neurodevelopmental sample, where they observed differences between sexes in relation to how memories are encoded in the hippocampus. In another neurodevelopmental study by Hwang, Velanova, and Luna (2010), the authors observed that improvements in inhibitory control were linked to strengthening of top-down connectivity for regions implicated in cognitive control networks, while similar findings were found in relation to top-down processing for a language network (Bitan et al., 2006). Further, it has been shown that there are differences in directionality in adolescent boys with externalizing behavior disorders in comparison with controls (Shannon, Sauder, Beauchaine, & Gatzke-Kopp, 2009). Likewise, alterations in connectivity direction in resting-state fMRI (rsfMRI) data have been observed for clinical samples, where children with autism and Attention Deficit Hyperactivity Disorder (ADHD) show a common pattern implicating Broca’s area across clinical groups, along with distinctive patterns of functional connectivity specific for given disorders (Henry et al., 2019). In addition, others have found higher directed connectivity from left to right superior parietal lobule to be associated with an improved shift and emotional control across healthy children and clinical participants (Dajani et al., 2019).

However, the literature on connection directionality is scarce, calling for validation of earlier findings and for new insights using novel approaches. In particular, we do not currently know how and to what extent the direction flow in the communication between brain networks is implicated in mental traits and disorders, including cognitive abilities. Given that cognitive deficits are seen across mental disorders, connecting these behavioral and clinical characteristics is crucial for gaining a better understanding of how higher-order processing is characterized in children and adolescence. Such integrative understanding is also important given that no consensus has been reached when it comes to the disorder-specific connectivity alterations that characterize particular mental disorders and the known overlap of symptoms (Craddock & Owen, 2010; Jollans & Whelan, 2018) and genetic pleiotropy between disorders (Lee et al., 2019; Smeland et al., 2019).

Dynamic graphical models (DGM) is a Bayesian approach for examining directed functional connectivity using dynamic linear models (DLM) (Schwab et al., 2018). As such, it implements a state space model that is linear and Gaussian in form. This includes statistically stationary properties as it uses a hidden Markov modelling approach, although it incorporates time varying coefficients and as a result can provide information about directionality in the form of a binary view of coupling between two brain regions for each connectivity direction.

Here, we used DGM and publicly available rsfMRI data from the Healthy Brain Network (HBN) project (Alexander et al., 2017) to study brain network information flow in children and adolescents and its associations with age, sex, cognitive abilities and psychopathology. We hypothesized that information flow would have a positive association, and be strengthened with increasing age (Bitan et al., 2006; Hwang et al., 2010), that there would be differences between females and males in the maturation of brain networks and its information flow (Riley et al., 2018), as well as alterations associated with information flow for control networks as these nodes are central to a range of disorders (Cortese, Kelly, & Di Martino, 2012; Eisenberg & Berman, 2010; Francx et al., 2015; Geiger et al., 2016; Li et al., 2017; Zhao, Swati, Metmer, Sang, & Lu, 2019).

## Methods

### Study samples

The HBN is a project organized by the Child Mind Institute (Alexander et al., 2017) and is a resource targeting novel insight into the critical time period when psychiatric and mental disorders emerge. The HBN consortium aims to include 10,000 individuals in the age range of 5-21 years from the New York area, where participants are included by use of announcements that are distributed to community members, educators, local care providers with the addition of sending information via email lists and events to parents, encouraging participation of children with clinical concerns to this study (Alexander et al., 2017). Data on these individuals include a package consisting of MRI scanning, genetics, electroencephalography (EEG), eye-tracking, as well as biological testing and a neuropsychological battery consisting of cognitive, lifestyle indices, behavioral and psychiatric domains in addition to actigraphy and voice and video interviews (Alexander et al., 2017). Exclusion criteria include serious neurological disorders, neurodegenerative disorders, acute encephalopathy, hearing or visual impairment, lifetime substance abuse that necessitated chemical replacement therapy/acute intoxication at time of study, recent diagnosis of a severe mental disorder or manic/psychotic episode within the last 6 months without ongoing treatment, in addition to the onset of suicidality/homicidality where there is no current, ongoing treatment (Alexander et al., 2017). All participants over the age of 18 years provided signed informed consent, while legal guardians signed informed consent for participants under the age of 18, in addition to participants giving a written assent (Alexander et al., 2017). The Chesapeake Institutional Review Board approved the study (https://www.chesapeakeirb.com/).

### MRI acquisition and preprocessing

MR data was collected by the study team of HBN, where we included MRI data from the following sites; Rutgers University Brain Imaging Center (RUBIC), Citigroup Biomedical Imaging Center (CBIC) and a mobile scanner located in Staten Island. MRI data was collected using one scanner at each site, giving a total of 3 scanners comprising our sample. Rutgers applied a Siemens 3T Tim Trio scanner, while CBIC utilized a Siemens 3T Prisma, and both sites applied the same MRI parameters, where resting-state blood-oxygen-level-dependent (BOLD) fMRI data was collected for each subject using a T2*-weighted BOLD echo-planar imaging (EPI) sequence with a repetition time (TR) of 800ms, echo time (TE) of 30ms, multiband acceleration factor = 6, 60 number of slices with the rsfMRI session consisting of 375 volumes and with voxel size= 2.4×2.4×2.4 mm. The third mobile scanner located in Staten Island used a 1.5T Siemens Avanto system equipped with 45 mT/m gradients (Alexander et al., 2017), where the following parameters were implemented; TR= 1.45s, TE=40ms, multiband acceleration factor = 3, number of volumes=420, slices= 54, resolution in mm= 2.5×2.5×2.5 mm (for more information about the MRI parameters, see http://fcon_1000.projects.nitrc.org/indi/cmi_healthy_brain_network/MRI%20Protocol.html). Raw imaging data was downloaded from the HBN database (http://fcon_1000.projects.nitrc.org/indi/cmi_healthy_brain_network/sharing_neuro.html#Direct%20Down), and analyzed on the secure data storage and computing facilities (TSD, https://www.uio.no/tjenester/it/forskning/sensitiv) at University of Oslo. Image processing tools, based largely on Smith et al. (2013), were used for the functional data while T1-weighted data, which was applied as an intermediate in the registration, was processed using FreeSurfer 5.3 (http://freesurfer.net), including removal of non-brain tissue.

As part of the HBN MRI protocol, multiple T1weighted sequences were acquired for each subject, and we used MRIQC version 0.14.2 (Esteban et al., 2017) for automated quality assessment (N=2427, CUNY scanning site was included for the MRIQC stage, yet this site was dropped at a later stage due the low sample size, N=22). From this, T1 weighted images with the best image quality metrics, based on the classifier ratings, were used as input for registration, while structural scans that were flagged as low quality (classifier rating score >0.5, N=160) were manually checked and excluded (N=117). In addition, 22 subjects had errors when they were run through FreeSurfer and after manually inspecting these datasets, these were also omitted due to motion artefacts and/or having MRI findings disrupting the segmentation pipeline in FreeSurfer. We also made a global mask of the T1 data where we manually checked subjects that had low coverage, excluding another 95 subjects leaving a total of 2171 datasets that had a usable T1w sequence for registration.

Two of the MRI sites, RUBIC and CBIC, had two resting-state scans acquired as part of the same MRI session. As the DGM method benefits from more time points, we merged the time series for subjects with two resting state scans together to leverage all available resting-state data. This was done prior to implementing feat as a means of optimizing the data by improving the spatial alignment between the sessions, and to have the full set of volumes to inform FIX.

Furthermore, the first five volumes for the fMRI dataset were discarded. We preprocessed fMRI data using FSL 6.0.3 (http://fsl.fmrib.ox.ac.uk), including motion correction and brain extraction. FSL’s FEAT (Woolrich, Ripley, Brady, & Smith, 2001) included spatial smoothing with a Gaussian kernel FWHM of 6 mm and a high-pass filter cutoff of 100. FMRIB’s Nonlinear Image Registration tool (FNIRT) was used to register fMRI volumes to standard space (MNI-152) with the T1w volumes as intermediates.

We also implemented a cleaning step for fMRI data, where we made a global mask for the fMRI datasets (which indicated data coverage across participants), and from this we excluded 128 of 1813 datasets with poor coverage. To reduce the influence of noise in the data and increase the tSNR (Kaufmann et al., 2017), we removed artefacts by use of ICA-AROMA, a classifier that identifies and removes motion specific noise in fMRI data (Pruim, Mennes, Buitelaar, & Beckmann, 2015; Pruim, Mennes, van Rooij, et al., 2015). Afterwards, ICA was rerun and FSL’s FIX (FMRIB’s ICA-based X-noisiefier (Griffanti et al., 2014; Salimi-Khorshidi et al., 2014)), was used with the recommended threshold of 20 to remove remaining motion confounds and other artefacts in the data. Further, data was temporally demeaned and variance normalized (Beckmann & Smith, 2004), and the quality controlled fMRI dataset (N=1685) was submitted to a group ICA, utilizing FSL’s Multivariate Exploratory Linear Optimized Decomposition into Independent Components (MELODIC) tool (Beckmann & Smith, 2004; Hyvärinen, 1999), where 25 components were extracted from the ICA and used for further analysis. Dual regression was applied to estimate individual spatial maps and corresponding time series from the group ICA (Beckmann & Smith, 2004; Filippini et al., 2009), which were used as input for the DGM analysis.

### Mental and cognitive measures

From the HBN data release, N=1685 participants were included after quality assessment. Out of these, information on age at MR was missing for N=45, sex for N=9, and cognitive/clinical information for 496 individuals. The participants were 5-22 years (mean: 11.5, years, sd: 3.51 years) and 37.9% were females. This data was used to study group-level patterns of dFC (average connectivity matrix). For the subsequent associations with age, sex, cognition and mental health, we restricted the analysis to a subset based on data availability. Thus, the final sample for the association analyses comprised N=1143, individuals aged 6-17 years (mean: 10.7 years, sd: 2.62 years, 37.4% females, where N=83 where from the Staten Island site, N=503 from CBIC and N=557 from RUBIC scanning site (see SI SFig.2 for age distributions within scanner sites). A large proportion of the sample had a diagnosis (N=1012), while N=105 did not have a diagnosis, N=23 dropped out before a diagnosis could be determined, and N=3 was missing information for clinical consensus diagnosis data and had as such not received a diagnosis. Broadly, the majority of patients included in our analysis had disorders in the following categories where comorbidities are included. Accordingly, there is a higher N than the total number of subjects for each diagnosis category given here: Neurodevelopmental disorders (N=1526), anxiety disorders (N=557), disruptive, impulsive control and conduct disorders (N=171), elimination disorders (N=111), and depressive disorders (N=115), see SI; SFig. 1 for further details.

We used the full-scale intelligence quotient (FSIQ) from the Wechsler Intelligence Scale for Children (WISC-V) taken for participants aged 6-17 years as a proxy for cognitive ability. This composite score includes the following domains; visual spatial, verbal comprehension, fluid reasoning, working memory, and processing speed (Wechsler, 2003). Mental health was measured on a continuum including both healthy subjects and patients as it is difficult to uncover robust findings for psychiatric diagnosis that constitutes heterogeneous disorders, showing a wide range in symptoms, severity, duration and prognosis, and as patients often have more than one diagnosis. Such heterogeneity is also reflected in the brain, making the search for biological markers a complex task. As such, for mental health, we performed a principal component analysis (PCA) on The Extended Strengths and Weaknesses Assessment of Normal Behavior (E-SWAN), which has been shown to be a valid psychometric assessment tool for investigating behavior underlying DSM disorders (Alexander, Salum, Swanson, & Milham, 2020). E-SWAN domains include depression, social anxiety, disruptive mood dysregulation disorder (DMDD), and panic disorder. We excluded 3 of the items relating to panic disorder from the questionnaire that had a high degree (90%) of missing values, giving a total of 62 items for the PCA analysis. The remaining items had available data for 2626 participants with no missing values. We performed PCA using the “prcomp” function in R, where the first PC, often denoted as the p-factor or pF (Caspi et al., 2013), explained 43.6% of the variance (Figure 1). From the loadings from the PCA, this component was associated with items related to self-control and depression/anxiety (Figure 1). In accordance with other studies showing more than one factor being of importance (Alnaes et al., 2018; Mallard et al., 2019), we also included the second principal component referred to as pF_2,_ which was associated with items relating to mood dysregulation. This component explained 11.3% of the variance.

**Figure 1:**
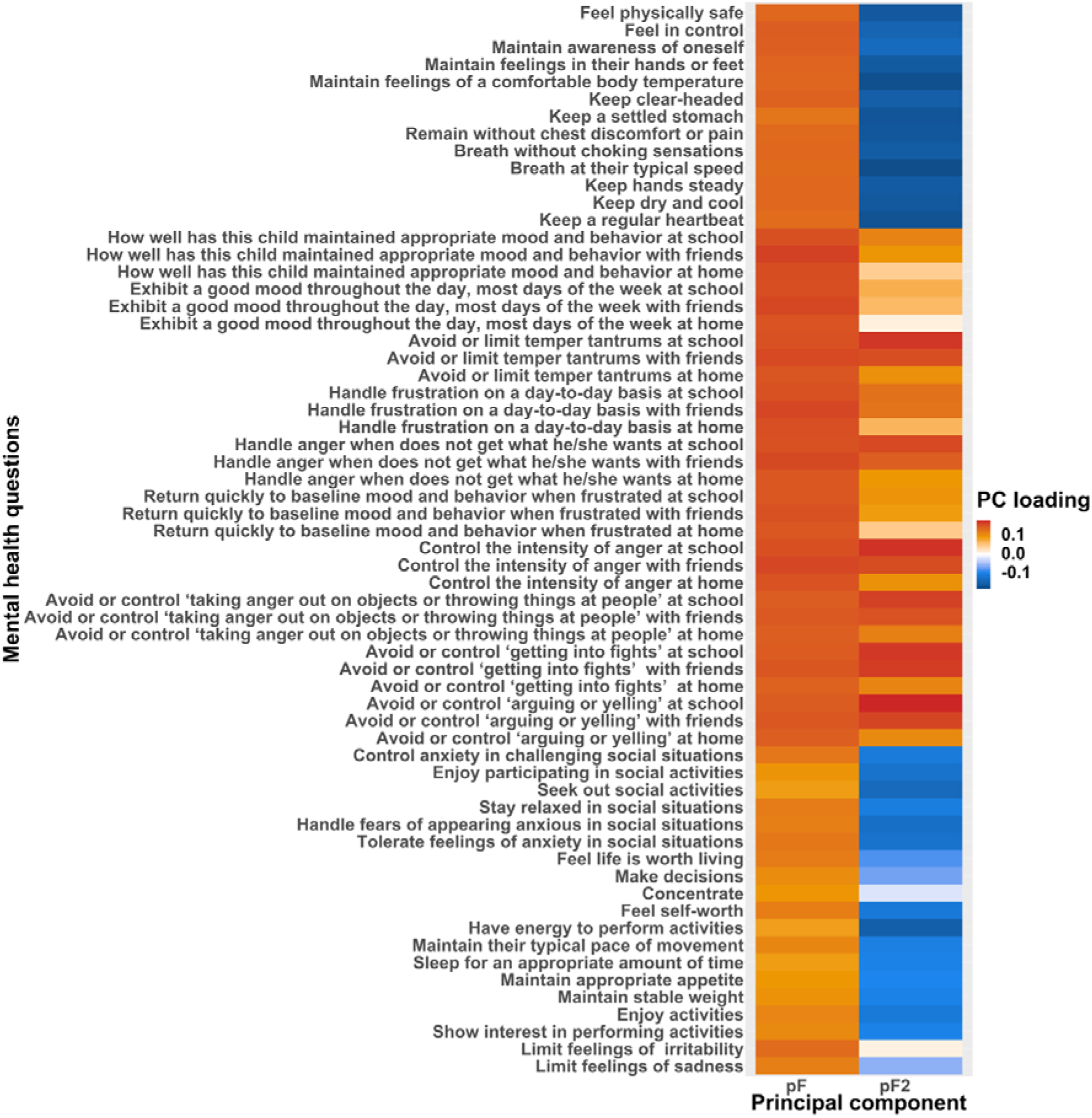
Principal component analysis performed on the ESWAN questionnaire taken as part of the HBN protocol. For visualization purposes, only the components included in our analysis are shown. We used the first two principal components as proxies of psychopathology, referred to as “pF” and “pF_2_”.

### Network analysis

We chose ten resting-state networks from the model order of 25 for inclusion in the dFC analysis. These networks were chosen based on spatial correlation and manually ensuring overlapping ICs with the ten RSNs reported by Smith et al. (2009). The ten networks comprised the default mode (DMN), cerebellar (Cer), visual occipital (VO), visual medial (VM), visual lateral (VL), right frontoparietal (FPR), left frontoparietal (FPL), sensorimotor (SM), auditory (Au), and the executive control (Ex) network (Figure 2). These networks were also used in previous DGM studies (Lund et al., 2020; Schwab et al., 2018). Including the same nodes allowed us to compare results with prior findings and to integrate the current results obtained from a childhood and adolescent sample with previous adult studies. The included timeseries for each node were mean centered before estimating dFC from individual level RSN time series using the DGM package v1.7.2 in R. DGM is a set of regressions with time-varying coefficients (DLMs) for every receiving node. The receiver node refers to the network that receives information from a sending node that transmits information between a node pair. For each DLM, all possible models for the sender nodes are tested and the model with the highest model evidence is selected. Consequently, a model is stated by the sending networks of a node, giving a binary outcome. The outcome indicates if there is an influence or not between two networks. DGM has uncovered meaningful trajectories in rsfMRI mice data where DGM revealed a link between areas in the hippocampus that fed information to the cingulate cortex, in line with previous studies using viral tracers to delineate directed anatomical connectivity in mice (Schwab et al., 2018). Additionally, even with systemic hemodynamic lag confounds being introduced in the data, network simulations demonstrated a sensitivity of 72%–77% for the DGM approach (Schwab et al., 2018).

**Figure 2:**
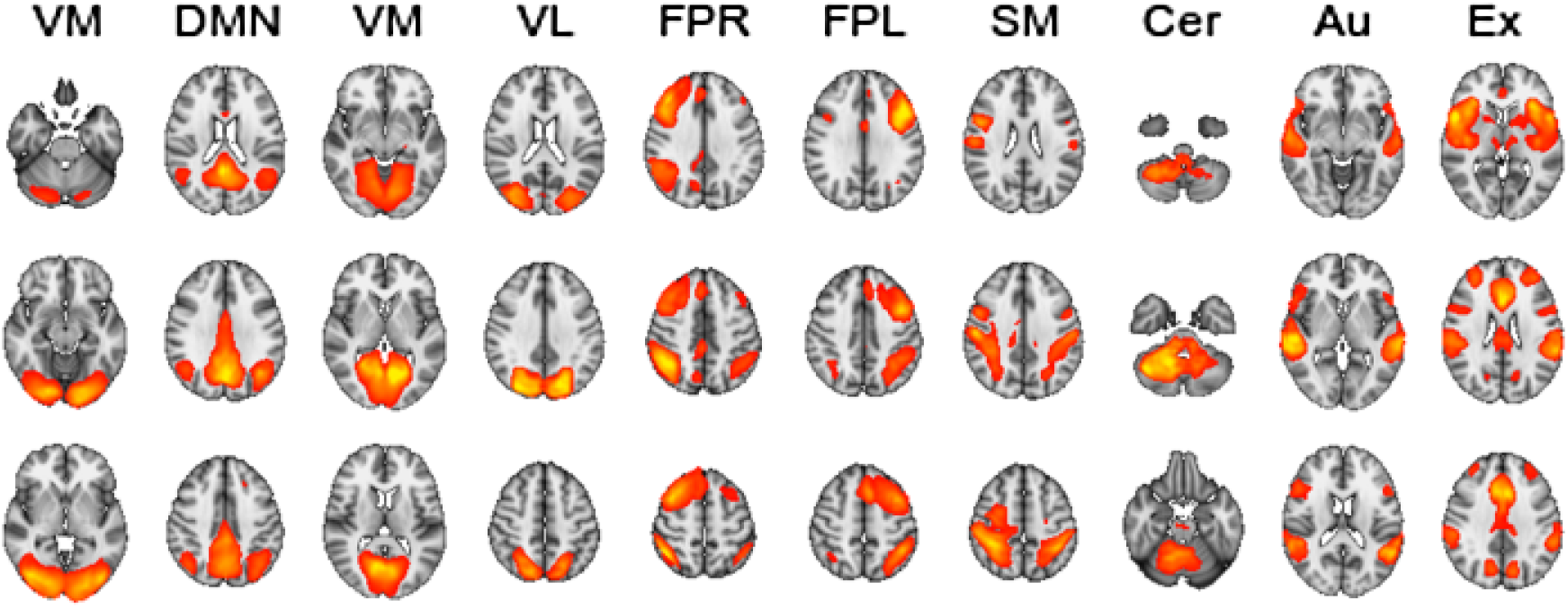
group-level maps for the 10 selected independent components included in the analysis (where the z-score maps threshold is set at 4).

### Statistical analysis

We included all available rsfMRI scans for the group ICA, as a higher number of subjects is beneficial for yielding more robust ICs, while restricting the association analyses to the subset that had all covariates available. We performed the same analysis as previously reported in an adult sample (Lund et al., 2020), examining dFC on the edge- and node-level. Edge-level analysis deployed logistic regression for every connection of the directed network using directed connectivity as the response variable and testing for associations with age, age-orthogonalized age squared (age^2^, using the poly function in R), sex, cognitive abilities, mental health, tSNR, motion and scanning site (as data was acquired at multiple scanners). These covariates were included into one model (see SI; SFig.5, for additional analyses examining potential multicollinearity for covariates included in the model). P-values were Bonferroni corrected for a number of 90 analyses on the edge-level (alpha level of 0.05). In addition, we performed node-level analysis to examine which networks overall send and receive information to each other (Lund et al., 2020). We calculated the number of output connections or outgoing edges (referred to as out-degree) and the number of input connections or incoming edges (referred to as in-degree) for a given node. We performed linear regression using in-degree and out-degree as dependent variables and the same independent variables as used in the logistic regression on edge-level. P-values were Bonferroni corrected for a number of 10 analysis on the node-level (alpha level of 0.05).

## Results

Figure 3 depicts the average directed functional connectivity matrix across all individuals. Several connections indicated bi-directionality in information flow, especially directionality estimates for the visual lateral and visual medial networks where information flow from VM to VL was present in 94.4% of individuals, while connection from VL to VM was present in 94.2%. In addition, there was overall a high number of participants that showed a reciprocal relationship between the VO to VM and VL (VO to VL: 89.1%, VO to VM: 80.2%) and opposite (VL to VO: 91.5%, VM to VO: 82.7%). These bi-directional relationships are in alignment with patterns of information flow found in adults from the UK Biobank sample (Lund et al., 2020). However, we did not observe the bi-directional information flow found previously for adults between the DMN and FPR network. Further, when observing receiver networks, the FPL showed the same as found for adults, that it did not receive information to a large extent (Lund et al., 2020), coherent with our findings in youths. Additionally, the edge most present across all individuals, aside from the visual networks, was Cer which receives input from the Au (85.6%). This connection was also relatively strongly expressed in the other direction (68.6%). Moreover, the FPL sent information to Cer, and this edge was present in 77.7% of individuals. Yet, in this neurodevelopmental sample we did not observe that Cer and Au overall were mostly receivers, as found for adults (Lund et al., 2020).

**Figure 3:**
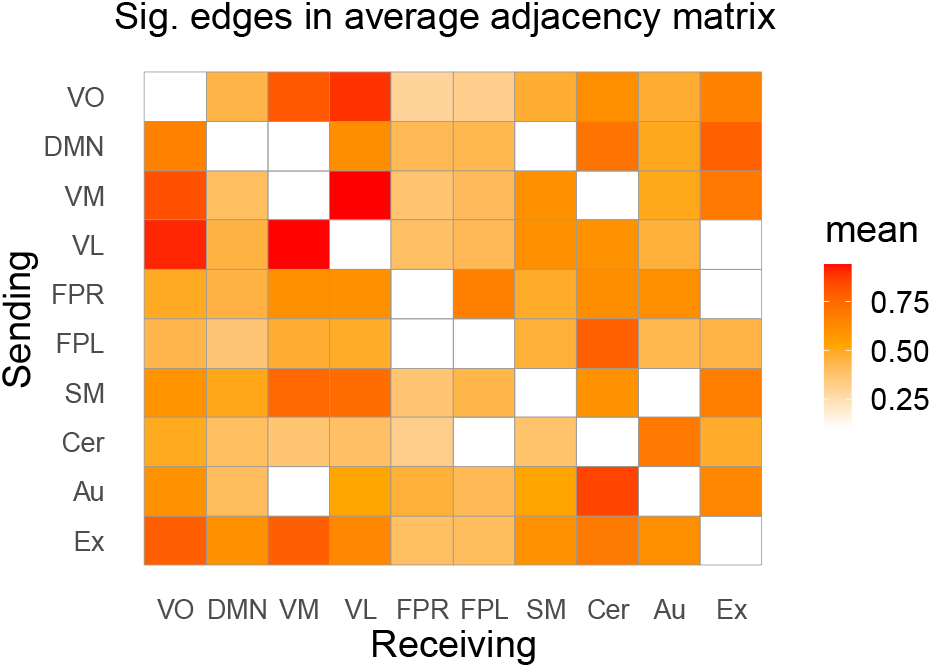
Average directed connectivity matrix across HBN subjects showing the significant proportions of edges between the 10 RSNs included in the analysis; VO, visual occipital; DMN, default mode; VM, visual medial; VL, visual lateral; FPR, frontoparietal right; FPL, frontoparietal left; SM, sensorimotor; Cer, cerebellum; Au, auditory; Ex, executive control network. The y-axis indicates the sender node, and the x-axis indicates the receiver nodes. Significance was assessed using a binomial test as implemented in the binom.nettest function in R, with a predefined FDR threshold of 5% and the hypothesized probability p0 = .56.

### Effects of age and cognition on dFC measures

Edge-level analysis of dFC showed significant effects of age, in line with our hypothesis of age being associated with the functional networks’ maturation processes occurring in childhood and adolescence (Figure 4; SI Tables 1-6 show z-scores and p-values and SFig.3-4 show effects of scanner). Specifically, we observed a positive association with age and dFC from the Cer to the Au network. Correspondingly, the Au sends more information with higher age to the SM node. For cognitive test performance and dFC, we observed a significant negative association between VO and DMN, with the DMN receiving less information from the VO network with higher cognitive test performance.

**Figure 4:**
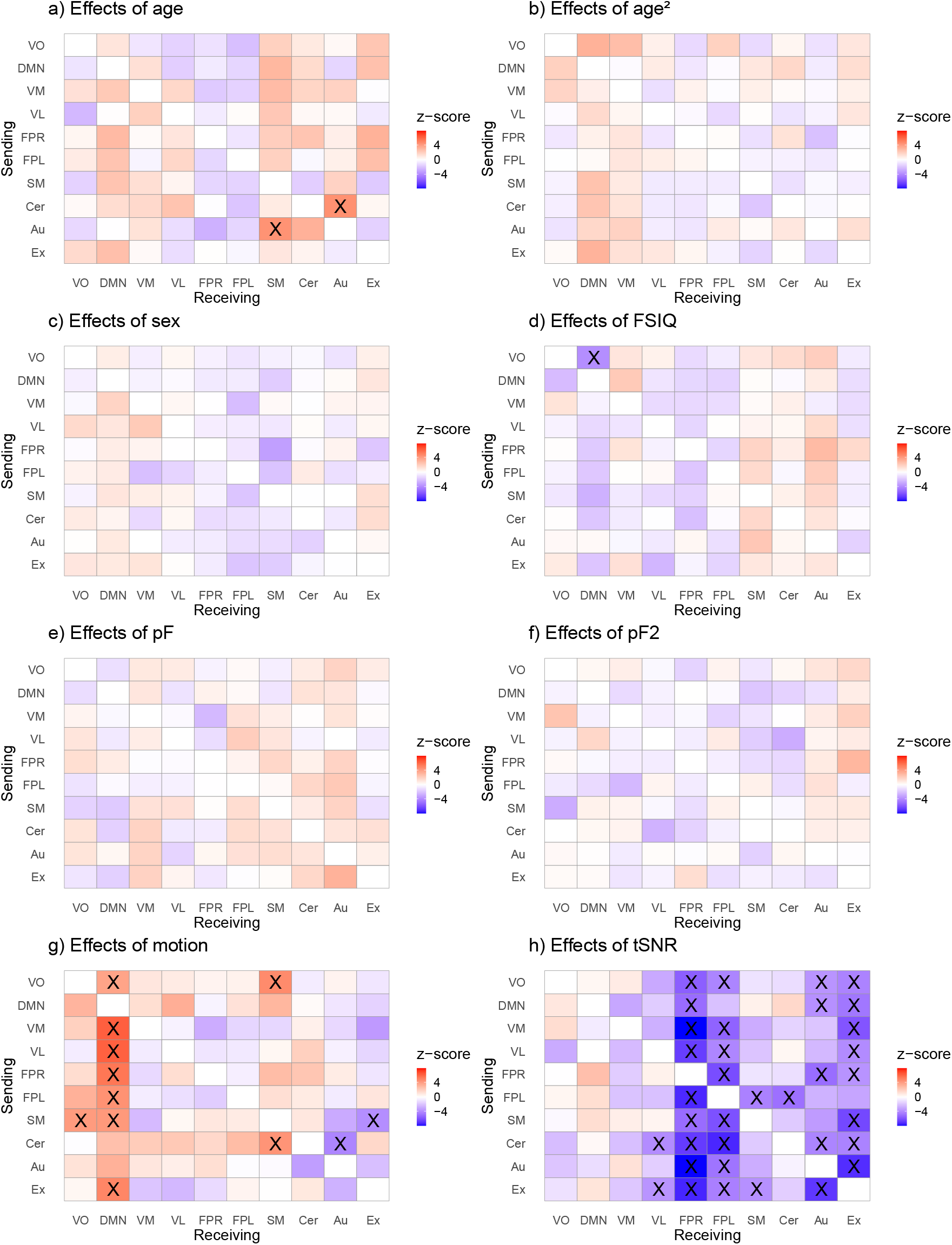
Matrices showing the effects of age (a), age^2^ (b), sex (c), intellectual abilities (FSIQ) (d), mental health (pF and pF_2_) (e,f), motion (g), and tSNR (h) on directed connectivity. The analysis was performed in HBN data that had no missing values (N = 1143, 6–17 years, df = 1132). Significant edges following Bonferroni correction are marked as X. The legend shows the 10 RSNs included in the analysis; VO, visual occipital; DMN, default mode; VM, visual medial; VL, visual lateral; FPR, frontoparietal right; FPL, frontoparietal left; SM, sensorimotor; Cer, cerebellum; Au, auditory; Ex, executive control network. The y-axis indicates the sender node, while the x-axis refers to the receiving node. The colors reflect the z-value for the corresponding effects where red indicates a positive association and blue a negative association.

**Figure 5:**
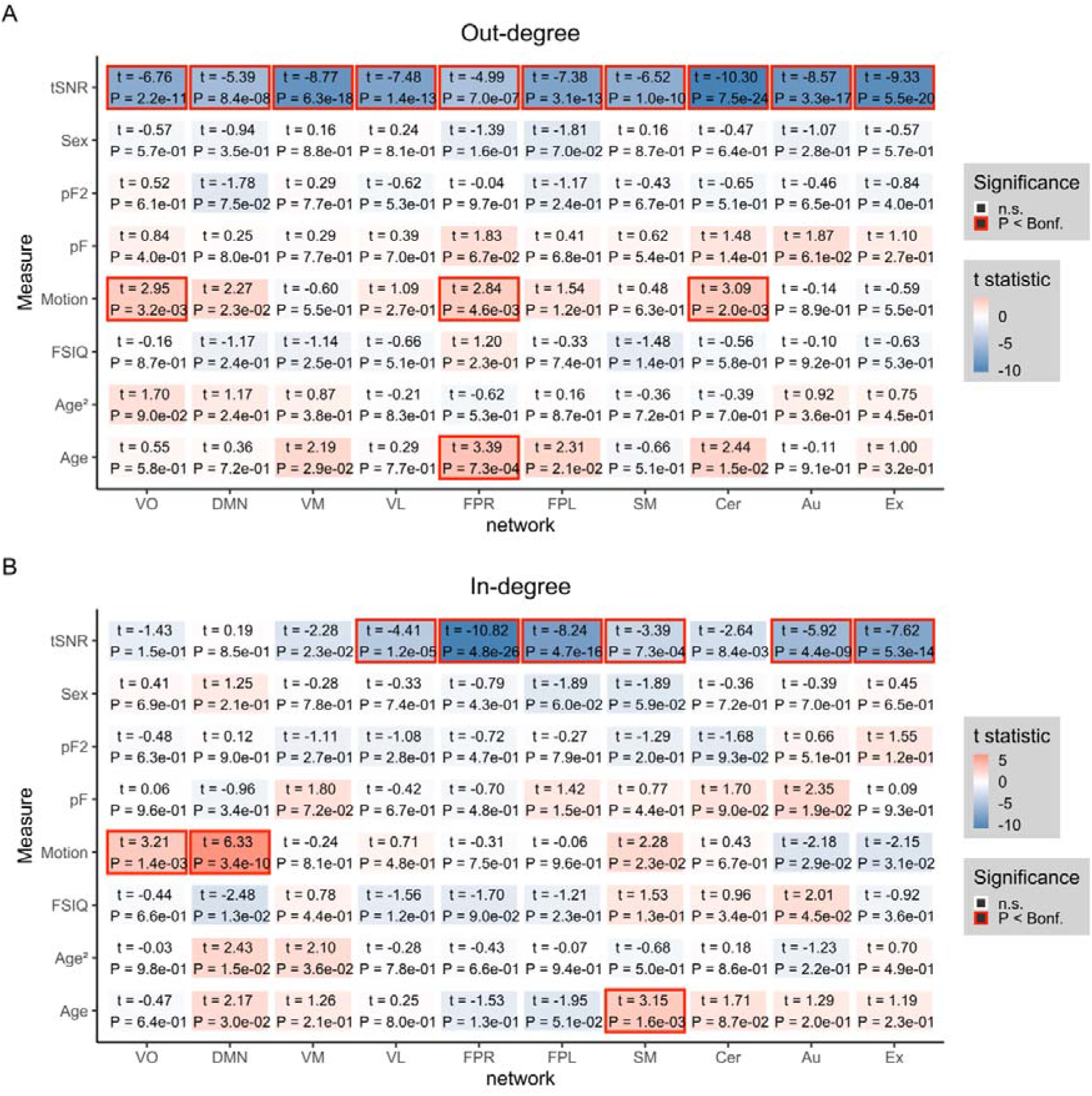
Associations on the node level (N=1143, 6-17 years, df= 1132). a) Out-degree matrix with corresponding effects of covariates age, age^2^, sex, cognition (FSIQ), mental health (pF and pF_2_), tSNR, and motion in HBN data. B) In-degree matrix with corresponding effects for the same covariates as in panel a). The colors reflect the t-value for the corresponding effect where numbers inside the boxes indicate t-statistic and p-value, and significant effects are marked with a red border following Bonferroni correction (p < .0.05).

On the node-level, we assessed similar phenotypic associations for the out-degree and in-degree of the networks. Confirming edge-level results, age was significantly associated with the in-degree (number of input connections) of SM (fig.4b), indicating that the SM network overall receive more information with increasing age. Additionally, for out-degree (number of output connections), there was a significant positive association for age and FPR, signifying that the FPR network overall sends more information with increasing age. As expected, there was significant effects for the control variables scanning site (SFig.3-4), tSNR and motion on edge and node-level.

Based on prior studies, we expected that mental health would be associated with information flow for control networks, however, this was not observed. We also expected sex-related differences in the maturation of brain networks, but this was not found.

## Discussion

Our analysis revealed marked dFC patterns for the visual networks. Specifically, we observed a reciprocal information flow between the VM and VL in the investigated sample of children and adolescence. This is in alignment with previous findings in adults (Lund et al., 2020; Schwab et al., 2018), and coherent with primary sensory and motor cortical regions maturing first, and being present early on in children (Casey, Tottenham, Liston, & Durston, 2005). In contrast, we did not find that the cerebellum or auditory networks are mostly receivers of information as observed in adults (Lund et al., 2020; Schwab et al., 2018). However, apart from the visual circuits, the Cer received most information overall from the other transmitter networks, potentially indicating its role as a receiver, as found for adults.

We observed significant associations between age and directed connectivity both at edge and node-level. At the edge-level and with increasing age, the Cer had higher information flow to the Au network, while the Au sent more information to the SM network. Of note, these connections demonstrating age-dependent dFC have been implicated in neurodevelopmental disorders such as autism, which is characterized by symptoms related to hyper- and hypo sensitivity in relation to sensory modalities (Marco, Hinkley, Hill, & Nagarajan, 2011). Moreover, it has been reported that the cerebellar network is mostly a receiver of information from other attention and memory related regions in children with ADHD when estimating directed connectivity in rsfMRI using a seed-based approach (Zhao, Zheng, Yang, & Tian, 2017). The average connectivity across participants indicated that the FPL sent information to Cer in 77.7% of the participants, which is in line with the aforementioned findings. Hence, this could be an interesting avenue to investigate in relation to potential biomarkers in future studies.

Likewise, on the node-level, the SM received more information with increasing age, while the FPR sent more information with higher age to the other networks overall. These results indicate that directionality estimates in both bottom-up and top-down processing are involved in the transitional phase from child to adulthood across patients and healthy individuals. This is in line with prior findings implicating these networks in normal functional development (Muetzel et al., 2016). Similarly, alterations in the right frontoparietal network has been associated with ADHD, where children and adolescent ADHD combined type patients show a reduction in FPR regions, when probing spatial working memory by use of task-fMRI (Silk et al., 2005; Vance et al., 2007).

For cognitive abilities and dFC, we observed a significant negative association between VO and DMN, with the DMN receiving less information from the VO network with higher cognitive test performance. This could indicate that individuals that were more focused on the task at hand benefited from less information flowing from VO to DMN, potentially facilitating effective whole-brain connectivity which has been linked to cognitive efficiency (van den Heuvel, Stam, Kahn, & Hulshoff Pol, 2009). However, it is challenging to delineate information flow as this could be disrupted for many connections and give rise to different alterations in connectivity. Nonetheless we did expect to observe associations for control networks in relation to cognitive abilities as this has been reported by others (Cole, Yarkoni, Repovs, Anticevic, & Braver, 2012; Li & Tian, 2014; R. Li, Zhang, Wu, Wen, & Han, 2020). Interestingly, the core executive function cognitive flexibility, has in healthy adults been associated with strong within-system connectivity in higher order systems and from these networks to primary-sensory motor systems when estimating dFC (Chen et al., 2019). Altered within network connectivity in relation to cognitive flexibility and in general has also been reported for neurodevelopmental disorders (Dajani et al., 2019; Henry et al., 2019). Here we estimated information flow between networks with different functions rather than within-system connectivity (with the exception of the visual networks). In addition, we used a normed cognitive composite test score as a proxy for cognition. However, this may not be an optimal approach as test norms could have limitations when it comes to representativeness and number of subjects included in the normative sample, making it less sensitive for clinical populations and for the lowest scoring percentile.

We utilized two general p factors for measuring symptoms in this sample. These psychopathology loadings, were related to self-control and depression/anxiety (pF) and items pertaining to mood dysregulation (pF_2_). Top-down control functions for cortico-limbic regions have been linked to modifications for symptoms relating to mood dysregulation in development. For instance, it has been shown that increased centromedial amygdala-rostral anterior cingulate cortex functional connectivity is related to a higher level of anxiety and depression symptoms during early adulthood, while increased structural connectivity in centromedial amygdala-anterior ventromedial prefrontal cortex white matter was linked to augmented symptoms of anxiety and depression during late childhood (Jalbrzikowski et al., 2017). We did not observe significant associations between dFC and symptom burden for any of the networks. Our dimensional approach of estimating PCA on symptom data represents both healthy subjects and individuals with a psychiatric disorder on a symptom continuum rather than as cases and control. Such a dimensional approach may better capture intermediate phenotypes which map to brain biology and the neuronal mechanisms underlying the symptoms, compared to diagnostic categories (Hengartner & Lehmann, 2017; Krueger & Bezdjian, 2009). While the lack of significant associations in our study indicates that between-network dFC in large-scale brain networks is not sensitive to mental health in this sample, future research will need to investigate dFC in distinct subnetworks and by different symptom measures, and also consider structural connectivity.

We expected to find differences between sexes in relation to directed connectivity as this has been found previously in adolescence (Riley et al., 2018) using task-fMRI, and when examining dFC in adults (Lund et al., 2020). However, we did not find any significant sex differences in dFC on edge-or node-level in rsfMRI. This may partly relate to sample characteristics such as the uneven sex distribution and the inclusion of patients in our sample.

### Limitations

As with any MRI study, participant motion during scanning may confound the results, especially as it is known that both young individuals and those with a psychiatric diagnosis on average move more (Satterthwaite et al., 2012). We therefore implemented a stringent correction pipeline, where we implemented several steps of ICA and machine learning based cleaning (FIX and AROMA). We also included the mean relative motion measure from FSL as a covariate in our analysis step. Another relevant confound effect that could potentially bias our results and the comparison to previous work is variability in the ICA decompositions. We performed ICA in the study sample and selected components based on similarity to components reported by Smith et al. (2009). While this approach yielded overall good comparison to previous work, subtle differences between decompositions may retain and might impact the derived dFC patterns. For instance, sensorimotor nodes were divided into three independent components where the node with the best spatial overlap with the SM component in Smith et al. (2009) was chosen. Most likely including another sensorimotor related node would change the results of our analysis. Hence, our results call for follow-up analysis in more fine-grained networks. In this framework, different ways to parcellate the brain could be tested.

The majority of the individuals in the sample were diagnosed with a child and adolescence mental disorder. This was beneficial when examining associations of psychopathology on directionality estimates as it yielded a high number of patients in each group, but makes it difficult to delineate if average patterns of information flow are for typical or non-typical functional development. In addition, medication may have had an effect on our results, as participants were asked to discontinue stimulant medication but were still enrolled if they chose not to discontinue due to personal reasons or if recommended by their physician.

On a methodological note, DGM reflects instantaneous relationships and does not only look at lagged relationships, however sensitivity of DGM drops depending on the total offset between a node pair and can as such influence the estimation of these interactions. Further, DGM estimates binary connections, which may have reduced the sensitivity of the association analyses.

Lastly, whether differences in findings compared to previous studies in adults are of methodological origin, such as differences in preprocessing pipelines or network definitions, or can be explained by differences in dynamic connectivity between children and adults remains to be further investigated.

## Conclusions

To conclude, using a sample of children and adolescents, we showed that the direction of the information flow was age dependent for auditory, sensorimotor and cerebellar connections. In addition, the FPR had a higher degree of input connections with higher age. These findings contribute to the existing knowledge in the brain development field, and warrant further studies for replication in healthy samples as well as other clinical populations.

## Data Availability

The data incorporated in this work were gathered from the open access Healthy Brain Network resource. Software needed to estimate directed connectivity is available at https://github.com/schw4b/DGM.

http://www.healthybrainnetwork.org

https://github.com/schw4b/DGM

## Acknowledgements

This manuscript was prepared using a limited access dataset obtained from the Child Mind Institute Biobank, the HBN resource (http://www.healthybrainnetwork.org). Its initiatives are supported by philanthropic contributions from the following individuals, foundations and organizations: Margaret Bilotti; Brooklyn Nets; Agapi and Bruce Burkard; James Chang; Phyllis Green and Randolph Cōwen; Grieve Family Fund; Susan Miller and Byron Grote; Sarah and Geoff Gund; George Hall; Jonathan M. Harris Family Foundation; Joseph P. Healey; The Hearst Foundations; Eve and Ross Jaffe; Howard & Irene Levine Family Foundation; Rachael and Marshall Levine; George and Nitzia Logothetis; Christine and Richard Mack; Julie Minskoff; Valerie Mnuchin; Morgan Stanley Foundation; Amy and John Phelan; Roberts Family Foundation; Jim and Linda Robinson Foundation, Inc.; The Schaps Family; Zibby Schwarzman; Abigail Pogrebin and David Shapiro; Stavros Niarchos Foundation; Preethi Krishna and Ram Sundaram; Amy and John Weinberg; Donors to the 2013 Child Advocacy Award Dinner Auction; Donors to the 2012 Brant Art Auction.

This manuscript reflects the views of the authors and does not necessarily reflect the opinions or views of the Child Mind Institute.

This work was performed on the TSD (Tjeneste for Sensitive Data) facilities, owned by the University of Oslo, operated and developed by the TSD service group at the University of Oslo, IT-Department (USIT).

## References

Alexander, L. M., Escalera, J., Ai, L., Andreotti, C., Febre, K., Mangone, A.,… Milham, M. P. (2017). An open resource for transdiagnostic research in pediatric mental health and learning disorders. Scientific Data, 4(1). doi:10.1038/sdata.2017.181

Alexander, L. M., Salum, G. A., Swanson, J. M., & Milham, M. P. (2020). Measuring strengths and weaknesses in dimensional psychiatry. J Child Psychol Psychiatry, 61(1), 40-50. doi:10.1111/jcpp.13104

Alnaes, D., Kaufmann, T., Doan, N. T., Cordova-Palomera, A., Wang, Y., Bettella, F.,… Westlye, L. T. (2018). Association of Heritable Cognitive Ability and Psychopathology With White Matter Properties in Children and Adolescents. JAMA Psychiatry, 75(3), 287-295. doi:10.1001/jamapsychiatry.2017.4277

Beckmann, C. F., & Smith, S. M. (2004). Probabilistic Independent Component Analysis for Functional Magnetic Resonance Imaging. IEEE Transactions on Medical Imaging, 23(2), 137-152. doi:10.1109/tmi.2003.822821

Bitan, T., Burman, D. D., Lu, D., Cone, N. E., Gitelman, D. R., Mesulam, M. M., & Booth, J. R. (2006). Weaker top-down modulation from the left inferior frontal gyrus in children. NeuroImage, 33(3), 991-998. doi:10.1016/j.neuroimage.2006.07.007

Blakemore, S.-J. (2012). Imaging brain development: The adolescent brain. NeuroImage, 61(2), 397-406. doi:10.1016/j.neuroimage.2011.11.080

Casey, B. J., Tottenham, N., Liston, C., & Durston, S. (2005). Imaging the developing brain: what have we learned about cognitive development? Trends Cogn Sci, 9(3), 104-110. doi:10.1016/j.tics.2005.01.011

Caspi, A., Houts, R. M., Belsky, D. W., Goldman-Mellor, S. J., Harrington, H., Israel, S.,… Moffitt, T. E. (2013). The p Factor. Clinical Psychological Science, 2(2), 119-137. doi:10.1177/2167702613497473

Chen, O. Y., Cao, H., Reinen, J. M., Qian, T., Gou, J., Phan, H.,… Cannon, T. D. (2019). Resting-state brain information flow predicts cognitive flexibility in humans. Sci Rep, 9(1), 3879. doi:10.1038/s41598-019-40345-8

Cole, M. W., Yarkoni, T., Repovs, G., Anticevic, A., & Braver, T. S. (2012). Global connectivity of prefrontal cortex predicts cognitive control and intelligence. J Neurosci, 32(26), 8988-8999. doi:10.1523/JNEUROSCI.0536-12.2012

Connolly, C. G., Wu, J., Ho, T. C., Hoeft, F., Wolkowitz, O., Eisendrath, S.,… Yang, T. T. (2013). Resting-state functional connectivity of subgenual anterior cingulate cortex in depressed adolescents. Biol Psychiatry, 74(12), 898-907. doi:10.1016/j.biopsych.2013.05.036

Cortese, S., Kelly, C., Chabernaud, C., Proal, E.,, & Di Martino, A., Milham, M. P., Castellanos, F. X.. (2012). Toward systems neuroscience of ADHD: a meta-analysis of 55 fMRI studies.. American Journal of Psychiatry, 169(10), 1038–1055.

Craddock, N., & Owen, M. J. (2010). The Kraepelinian dichotomy - going, going… but still not gone. Br J Psychiatry, 196(2), 92-95. doi:10.1192/bjp.bp.109.073429

Dajani, D. R., Burrows, C. A., Nebel, M. B., Mostofsky, S. H., Gates, K. M., & Uddin, L. Q. (2019). Parsing Heterogeneity in Autism Spectrum Disorder and Attention-Deficit/Hyperactivity Disorder with Individual Connectome Mapping. Brain Connect, 9(9), 673-691. doi:10.1089/brain.2019.0669

Di Martino, A., Kelly, C., Grzadzinski, R., Zuo, X. N., Mennes, M., Mairena, M. A.,… Milham, M. P. (2011). Aberrant striatal functional connectivity in children with autism. Biol Psychiatry, 69(9), 847-856. doi:10.1016/j.biopsych.2010.10.029

Eisenberg, D. P., & Berman, K. F. (2010). Executive function, neural circuitry, and genetic mechanisms in schizophrenia. Neuropsychopharmacology, 35(1), 258-277. doi:10.1038/npp.2009.111

Esteban, O., Birman, D., Schaer, M., Koyejo, O. O., Poldrack, R. A., & Gorgolewski, K. J. (2017). MRIQC: Advancing the automatic prediction of image quality in MRI from unseen sites. PLoS One, 12(9), e0184661. doi:10.1371/journal.pone.0184661

Filippini, N., MacIntosh, B. J., Hough, M. G., Goodwin, G. M., Frisoni, G. B., Smith, S. M.,… Mackay, C. E. (2009). Distinct patterns of brain activity in young carriers of the APOE- 4 allele. Proceedings of the National Academy of Sciences, 106(17), 7209- 7214. doi:10.1073/pnas.0811879106

Francx, W., Oldehinkel, M., Oosterlaan, J., Heslenfeld, D., Hartman, C. A., Hoekstra, P. J.,… Mennes, M. (2015). The executive control network and symptomatic improvement in attention-deficit/hyperactivity disorder. Cortex, 73, 62-72. doi:10.1016/j.cortex.2015.08.012

Friston, K., Moran, R., & Seth, A. K. (2013). Analysing connectivity with Granger causality and dynamic causal modelling. Curr Opin Neurobiol, 23(2), 172-178. doi:10.1016/j.conb.2012.11.010

Friston, K. J., & Frith, C. D. (1995). Schizophrenia: a disconnection syndrome. Clin Neurosci, 3(2), 89–97.

Geiger, M. J., Domschke, K., Ipser, J., Hattingh, C., Baldwin, D. S., Lochner, C., & Stein, D. J. (2016). Altered executive control network resting-state connectivity in social anxiety disorder. World J Biol Psychiatry, 17(1), 47-57. doi:10.3109/15622975.2015.1083613

Griffanti, L., Salimi-Khorshidi, G., Beckmann, C. F., Auerbach, E. J., Douaud, G., Sexton, C. E.,… Smith, S. M. (2014). ICA-based artefact removal and accelerated fMRI acquisition for improved resting state network imaging. NeuroImage, 95, 232-247. doi:10.1016/j.neuroimage.2014.03.034

Hamm, L. L., Jacobs, R. H., Johnson, M. W., Fitzgerald, D. A., Fitzgerald, K. D., Langenecker, S. A.,… Phan K. L. (2014). Aberrant amygdala functional connectivity at rest in pediatric anxiety disorders. Biology of mood & anxiety disorders, 4(1)(15).

Hengartner, M. P., & Lehmann, S. N. (2017). Why Psychiatric Research Must Abandon Traditional Diagnostic Classification and Adopt a Fully Dimensional Scope: Two Solutions to a Persistent Problem. Front Psychiatry, 8, 101. doi:10.3389/fpsyt.2017.00101

Henry, T. R., Feczko, E., Cordova, M., Earl, E., Williams, S., Nigg, J. T.,… Gates, K. M. (2019). Comparing directed functional connectivity between groups with confirmatory subgrouping GIMME. NeuroImage, 188, 642-653. doi:10.1016/j.neuroimage.2018.12.040

Hoff, G. E., Van den Heuvel, M. P., Benders, M. J., Kersbergen, K. J., & De Vries, L. S. (2013). On development of functional brain connectivity in the young brain. Front Hum Neurosci, 7, 650. doi:10.3389/fnhum.2013.00650

Hwang, K., Velanova, K., & Luna, B. (2010). Strengthening of top-down frontal cognitive control networks underlying the development of inhibitory control: a functional magnetic resonance imaging effective connectivity study. J Neurosci, 30(46), 15535- 15545. doi:10.1523/JNEUROSCI.2825-10.2010

Hyvärinen, A. (1999). Fast and robust fixed-point algorithms for independent component analysis. IEEE Transactions on Neural Networks, 10(3), 626–634.

Insel, T. R. (2010). Rethinking schizophrenia. Nature, 468(7321), 187-193. doi:10.1038/nature09552

Jalbrzikowski, M., Larsen, B., Hallquist, M. N., Foran, W., Calabro, F., & Luna, B. (2017). Development of White Matter Microstructure and Intrinsic Functional Connectivity Between the Amygdala and Ventromedial Prefrontal Cortex: Associations With Anxiety and Depression. Biol Psychiatry, 82(7), 511-521. doi:10.1016/j.biopsych.2017.01.008

Jollans, L., & Whelan, R. (2018). Neuromarkers for Mental Disorders: Harnessing Population Neuroscience. Front Psychiatry, 9, 242. doi:10.3389/fpsyt.2018.00242

Kaufmann, T., Alnaes, D., Doan, N. T., Brandt, C. L., Andreassen, O. A., & Westlye, L. T. (2017). Delayed stabilization and individualization in connectome development are related to psychiatric disorders. Nat Neurosci, 20(4), 513-515. doi:10.1038/nn.4511

Keshavan, M. S., Giedd, J., Lau, J. Y. F., Lewis, D. A., & Paus, T. (2014). Changes in the adolescent brain and the pathophysiology of psychotic disorders. The Lancet Psychiatry, 1(7), 549-558. doi:10.1016/s2215-0366(14)00081-9

Kolskar, K. K., Alnaes, D., Kaufmann, T., Richard, G., Sanders, A. M., Ulrichsen, K. M.,… Westlye, L. T. (2018). Key Brain Network Nodes Show Differential Cognitive Relevance and Developmental Trajectories during Childhood and Adolescence. eNeuro, 5(4). doi:10.1523/ENEURO.0092-18.2018

Krueger, R. F., & Bezdjian, S. (2009). Enhancing research and treatment of mental disorders with dimensional concepts: toward DSM-V and ICD-11.. World Psychiatry, 8(1)(3).

Lee, P. H., Anttila, V., Won, H., Feng, Y.-C. A., Rosenthal, J., Zhu, Z.,… Smoller, J. W. (2019). Genomic Relationships, Novel Loci, and Pleiotropic Mechanisms across Eight Psychiatric Disorders. Cell, 179(7), 1469-1482.e1411. doi:10.1016/j.cell.2019.11.020

Li, C., & Tian, L. (2014). Association between resting-state coactivation in the parieto-frontal network and intelligence during late childhood and adolescence. AJNR Am J Neuroradiol, 35(6), 1150-1156. doi:10.3174/ajnr.A3850

Li, C. L., Deng, Y. J., He, Y. H., Zhai, H. C., & Jia, F. C. (2019). The development of brain functional connectivity networks revealed by resting-state functional magnetic resonance imaging. Neural Regen Res, 14(8), 1419-1429. doi:10.4103/1673-5374.253526

Li, P., Fan, T. T., Zhao, R. J., Han, Y., Shi, L., Sun, H. Q.,… Lu, L. (2017). Altered Brain Network Connectivity as a Potential Endophenotype of Schizophrenia. Sci Rep, 7(1), 5483. doi:10.1038/s41598-017-05774-3

Li, R., Zhang, J., Wu, X., Wen, X., & Han, B. (2020). Brain-wide resting-state connectivity regulation by the hippocampus and medial prefrontal cortex is associated with fluid intelligence. Brain Struct Funct, 225(5), 1587-1600. doi:10.1007/s00429-020-02077-8

Lund, M. J., Alnaes, D., Schwab, S., van der Meer, D., Andreassen, O. A., Westlye, L. T., & Kaufmann, T. (2020). Differences in directed functional brain connectivity related to age, sex and mental health. Hum Brain Mapp. doi:10.1002/hbm.25116

Mallard, T. T., Linnér, R. K., Okbay, A., Grotzinger, A. D., de Vlaming, R., Meddens, S. F. W.,… Harden, K. P. (2019). Not just one p: Multivariate GWAS of psychiatric disorders and their cardinal symptoms reveal two dimensions of cross-cutting genetic liabilities.. bioRxiv, 603134. doi:10.1101/603134

Marco, E. J., Hinkley, L. B., Hill, S. S., & Nagarajan, S. S. (2011). Sensory processing in autism: a review of neurophysiologic findings. Pediatr Res, 69(5 Pt 2), 48R-54R. doi:10.1203/PDR.0b013e3182130c54

Muetzel, R. L., Blanken, L. M., Thijssen, S., van der Lugt, A., Jaddoe, V. W., Verhulst, F. C.,… White, T. (2016). Resting-state networks in 6-to-10 year old children. Hum Brain Mapp, 37(12), 4286-4300. doi:10.1002/hbm.23309

Pruim, R. H. R., Mennes, M., Buitelaar, J. K., & Beckmann, C. F. (2015). Evaluation of ICA-AROMA and alternative strategies for motion artifact removal in resting state fMRI. NeuroImage, 112, 278-287. doi:10.1016/j.neuroimage.2015.02.063

Pruim, R. H. R., Mennes, M., van Rooij, D., Llera, A., Buitelaar, J. K., & Beckmann, C. F. (2015). ICA-AROMA: A robust ICA-based strategy for removing motion artifacts from fMRI data. NeuroImage, 112, 267-277. doi:10.1016/j.neuroimage.2015.02.064

Rausch, A., Zhang, W., Haak, K. V., Mennes, M., Hermans, E. J., van Oort, E.,… Groen, W. B. (2016). Altered functional connectivity of the amygdaloid input nuclei in adolescents and young adults with autism spectrum disorder: a resting state fMRI study. Mol Autism, 7, 13. doi:10.1186/s13229-015-0060-x

Riley, J. D., Chen, E. E., Winsell, J., Davis, E. P., Glynn, L. M., Baram, T. Z.,… Solodkin, A. (2018). Network specialization during adolescence: Hippocampal effective connectivity in boys and girls. NeuroImage, 175, 402-412. doi:10.1016/j.neuroimage.2018.04.013

Salimi-Khorshidi, G., Douaud, G., Beckmann, C. F., Glasser, M. F., Griffanti, L., & Smith, S. M. (2014). Automatic denoising of functional MRI data: combining independent component analysis and hierarchical fusion of classifiers. NeuroImage, 90, 449-468. doi:10.1016/j.neuroimage.2013.11.046

Satterthwaite, T. D., Wolf, D. H., Loughead, J., Ruparel, K., Elliott, M. A., Hakonarson, H.,… Gur, R. E. (2012). Impact of in-scanner head motion on multiple measures of functional connectivity: relevance for studies of neurodevelopment in youth. NeuroImage, 60(1), 623-632. doi:10.1016/j.neuroimage.2011.12.063

Schwab, S., Harbord, R., Zerbi, V., Elliott, L., Afyouni, S., Smith J. Q.,… Nichols T. E. (2018). Directed functional connectivity using dynamic graphical models. NeuroImage, 175, 340-353. doi:10.1016/j.neuroimage.2018.03.074

Schweinsburg, A. D., Nagel, B. J., & Tapert, S. F. (2005). fMRI reveals alteration of spatial working memory networks across adolescence. J Int Neuropsychol Soc, 11(5), 631-644. doi:10.1017/S1355617705050757

Shannon, K. E., Sauder, C., Beauchaine, T. P., & Gatzke-Kopp, L. M. (2009). Disrupted Effective Connectivity Between the Medial Frontal Cortex and the Caudate in Adolescent Boys With Externalizing Behavior Disorders. Criminal Justice and Behavior, 36(11), 1141-1157. doi:10.1177/0093854809342856

Silk, T., Vance, A., Rinehart, N., Egan, G., O’boyle, M., Bradshaw, J. L., & Cunnington, R. (2005). Fronto-parietal activation in attention-deficit hyperactivity disorder, combined type: functional magnetic resonance imaging study. The British Journal of Psychiatry, 187(3), 282–283.

Smeland, O. B., Frei, O., Fan, C.-C., Shadrin, A., Dale, A. M., & Andreassen, O. A. (2019). The emerging pattern of shared polygenic architecture of psychiatric disorders, conceptual and methodological challenges. Psychiatric Genetics, 29(5), 152-159. doi:10.1097/ypg.0000000000000234

Smith, S. M., Fox, P. T., Miller, K. L., Glahn, D. C., Fox, P. M., Mackay, C. E.,… Beckmann, C. F. (2009). Correspondence of the brain’s functional architecture during activation and rest. Proc Natl Acad Sci U S A, 106(31), 13040-13045. doi:10.1073/pnas.0905267106

van den Heuvel, M. P., Stam, C. J., Kahn, R. S., & Hulshoff Pol H. E. (2009). Efficiency of functional brain networks and intellectual performance. J Neurosci, 29(23), 7619-7624. doi:10.1523/JNEUROSCI.1443-09.2009

Vance, A., Silk, T. J., Casey, M., Rinehart, N. J., Bradshaw, J. L., Bellgrove, M. A., & Cunnington, R. (2007). Right parietal dysfunction in children with attention deficit hyperactivity disorder, combined type: a functional MRI study. Mol Psychiatry, 12(9), 826-832,793. doi:10.1038/sj.mp.4001999

Wechsler, D. (2003). Wechsler intelligence scale for children--Fourth Edition (WISC-IV)

Woolrich, M. W., Ripley, B. D., Brady, M., & Smith, S. M. (2001). Temporal autocorrelation in univariate linear modeling of FMRI data. NeuroImage, 14(6), 1370-1386. doi:10.1006/nimg.2001.0931

Zhao, D., Zheng, S., Yang, L., & Tian, Y. (2017). Causal connectivity abnormalities of regional homogeneity in children with attention deficit hyperactivity disorder: a rest-state fMRI study. ADMET and DMPK, 5(4), 242-252. doi:10.5599/admet.5.4.485

Zhao, Q., Swati, Z. N. K., Metmer, H., Sang, X., & Lu, J. (2019). Investigating executive control network and default mode network dysfunction in major depressive disorder. Neurosci Lett, 701, 154-161. doi:10.1016/j.neulet.2019.02.045

