## Supplementary Information for "Functional connectivity directionality between large-scale resting-state networks in children and adolescence from the Healthy Brain Network sample"

**Content**

*Page*

1. Diagnosis information	*1*

2. Age distributions within scanning sites	*2*

3. Scanner effects on edge- and node-level	*2*

4. Supplementary tables	*4*

5. Correlation among measurements	*7*

**1. Diagnosis information**

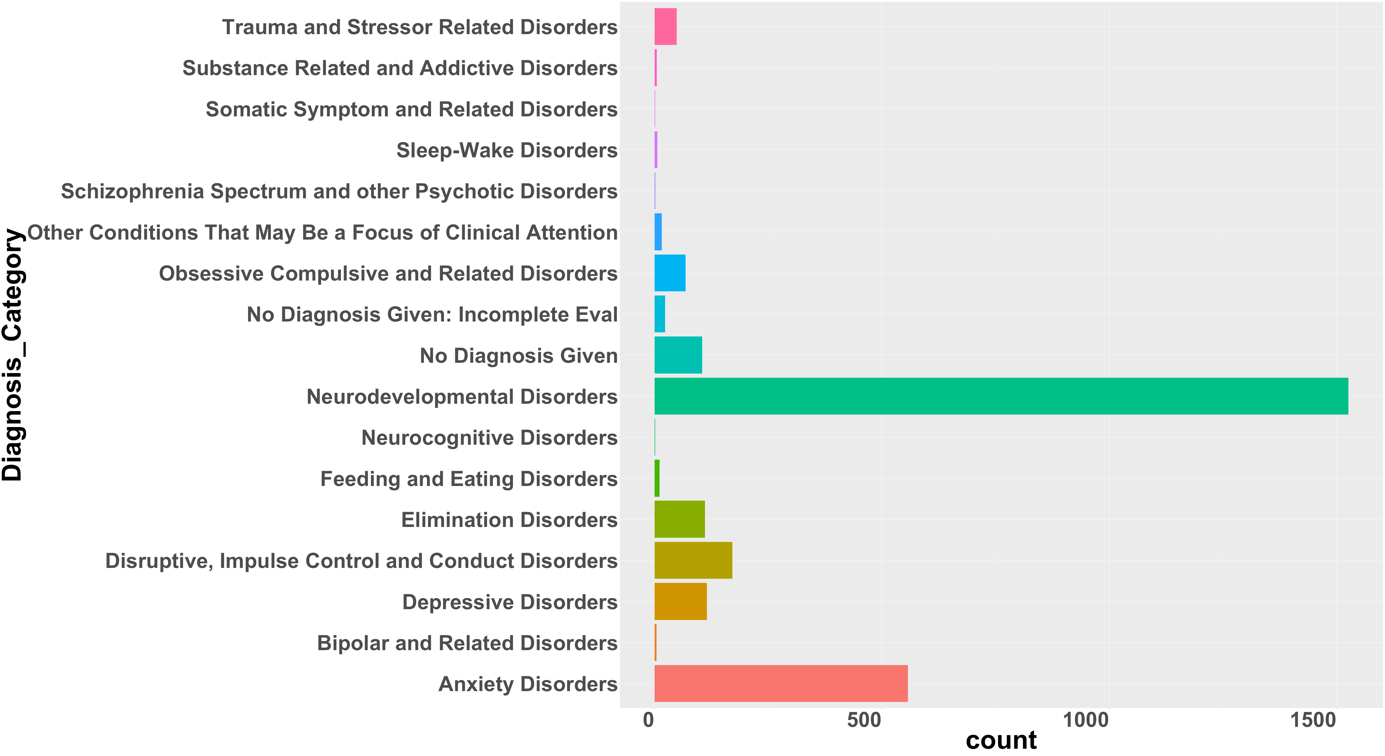

**Figure S1.** *Number of diagnoses given to HBN participants that were part of main analysis (N=1143), where diagnoses are grouped by category. This is based on the final consensus diagnosis given by the lead clinician at the end of participation.*

**2. Age distributions within scanning sites**

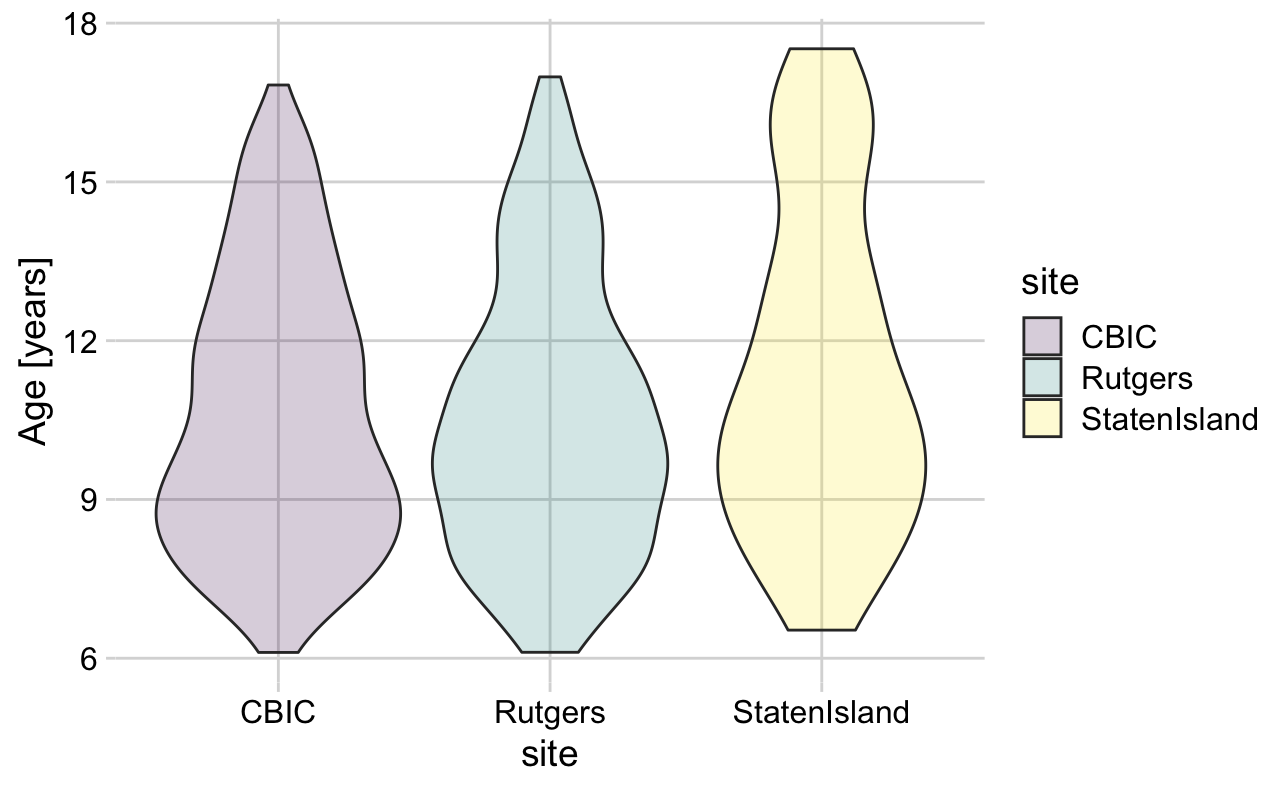

**Figure S2.** *Age distributions within scanning site for the HBN participants that were part of main analysis (N=1143), where N=83 was from the Staten Island site, N=503 from CBIC and N=557 from RUBIC/Rutgers scanning site.*

**3. Scanner effects on edge- and node-level**

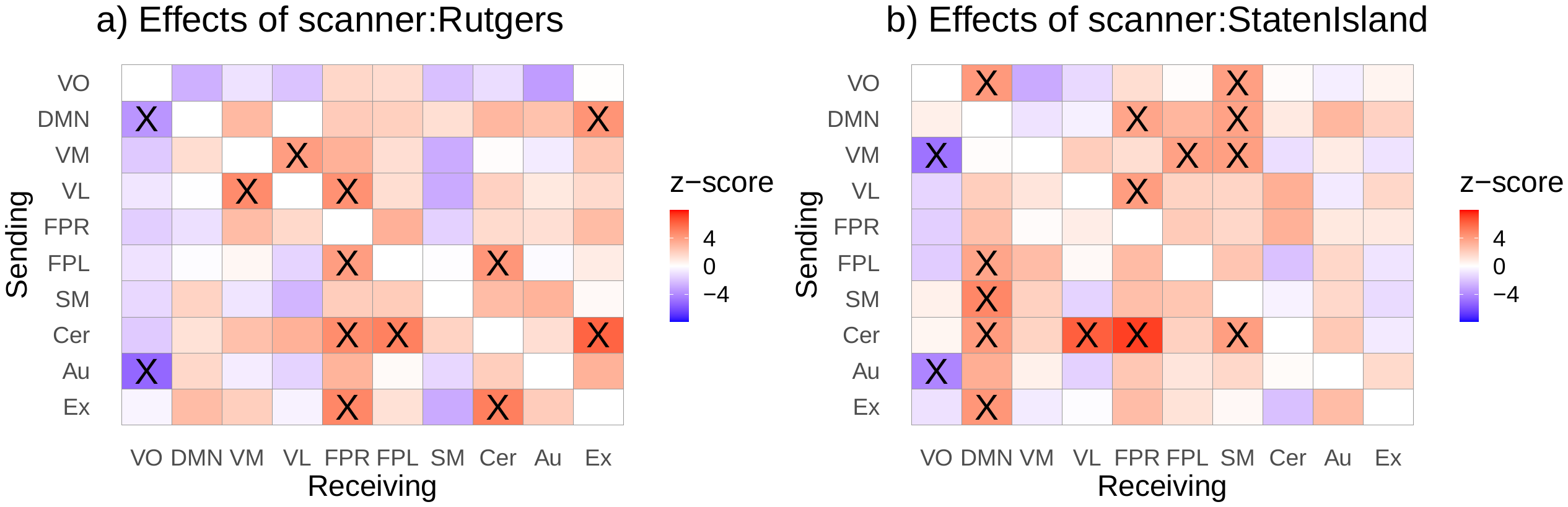

**Figure S3***: Matrices showing the effects of Rutgers scanner (a), and effects of scanner located at Staten Island (b). The analysis was performed in HBN data that had no missing values (N = 1143, 6–17 years, df = 1132). Significant edges following Bonferroni correction are marked as X. The legend shows the 10 RSNs included in the analysis; VO, visual occipital; DMN, default mode; VM, visual medial; VL, visual lateral; FPR, frontoparietal right; FPL, frontoparietal left; SM, sensorimotor; Cer, cerebellum; Au, auditory; Ex, executive control network. The y-axis indicates the sender node, while the x-axis refers to the receiving node. The colors reflect the z-value for the corresponding effects where red indicates a positive association and blue a negative association.*

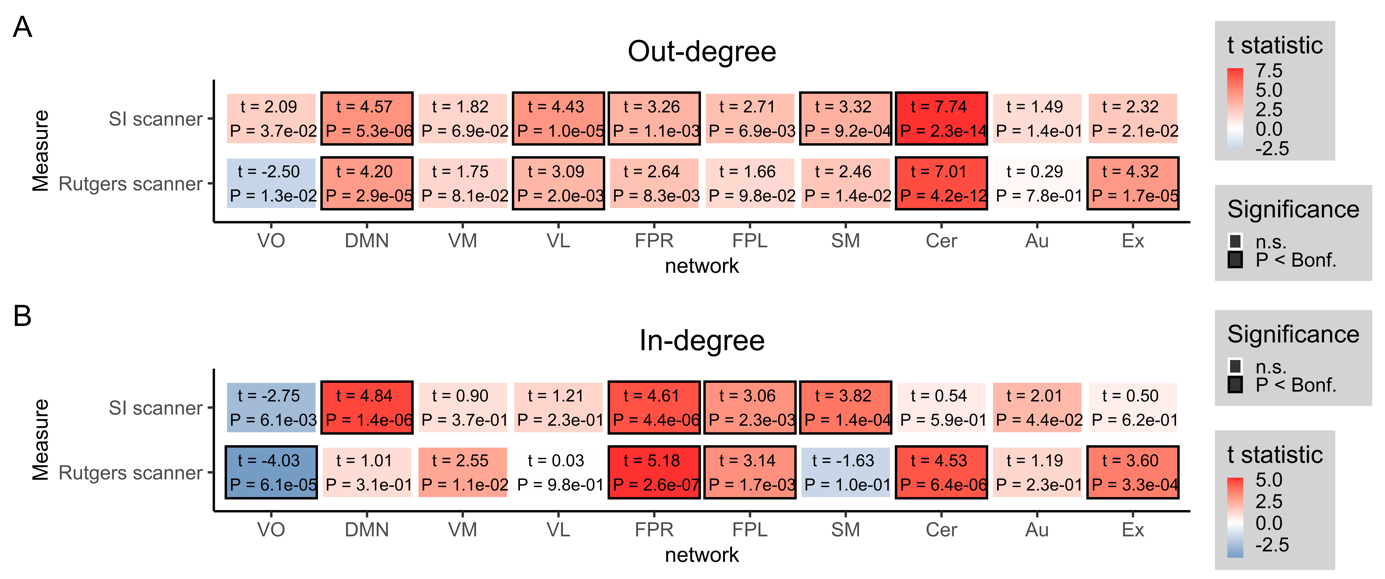

**Figure S4:** *Associations on the node level (N=1143, 6-17 years, df= 1132). a) Out-degree matrix with corresponding effects of covariates scanner in HBN data. B) In-degree matrix with corresponding effects for the same scanner covariates as in panel a). The colors reflect the t-value for the corresponding effect where numbers inside the boxes indicate t-statistic and p-value, and significant effects are marked with a black border following Bonferroni correction (p < .0.05).*

**4. Supplementary tables**

|  | **VO** | **DMN** | **VM** | **VL** | **FPR** | **FPL** | **SM** | **Cer** | **Au** | **Ex** |
| --- | --- | --- | --- | --- | --- | --- | --- | --- | --- | --- |
| **VO** | NA | 1.06, p =1 | -0.84, p =1 | -1.48, p =1 | -0.97, p =1 | -2.15, p =1 | 1.92, p =1 | 0.89, p =1 | 0.29, p =1 | 2.3, p =1 |
| **DMN** | -0.91, p =1 | NA | 1.25, p =1 | -1.63, p =1 | -0.71, p =1 | -1.38, p =1 | 2.91, p =0.32174 | 1.45, p =1 | -1.39, p =1 | 2.52, p =1 |
| **VM** | 1.23, p =1 | 2.21, p =1 | NA | 1.62, p =1 | -1.78, p =1 | -1.51, p =1 | 2.74, p =0.54927 | 1.71, p =1 | 1.85, p =1 | -0.06, p =1 |
| **VL** | -2.37, p =1 | 0.07, p =1 | 1.91, p =1 | NA | -0.63, p =1 | 0.13, p =1 | 2.27, p =1 | 0.27, p =1 | 0.23, p =1 | -0.71, p =1 |
| **FPR** | 0.39, p =1 | 2.81, p =0.43923 | 0.15, p =1 | 1.03, p =1 | NA | -0.86, p =1 | 2, p =1 | 2.38, p =1 | 0.68, p =1 | 3.18, p =0.13137 |
| **FPL** | 0.79, p =1 | 2.42, p =1 | -0.35, p =1 | 1.65, p =1 | -1.36, p =1 | NA | 1.89, p =1 | -0.25, p =1 | 0.63, p =1 | 2.62, p =0.78958 |
| **SM** | -1.51, p =1 | 2.42, p =1 | 1.29, p =1 | 0.43, p =1 | -1.37, p =1 | -1.45, p =1 | NA | -1.63, p =1 | 1.81, p =1 | -1.78, p =1 |
| **Cer** | 0.21, p =1 | 1.4, p =1 | 1.54, p =1 | 2.44, p =1 | -0.06, p =1 | -1.95, p =1 | 0.84, p =1 | NA | 4.45, p =0.00076 | 0.35, p =1 |
| **Au** | -1.26, p =1 | -0.02, p =1 | 1.14, p =1 | -1.14, p =1 | -2.64, p =0.75565 | -1.39, p =1 | 4.36, p =0.00118 | 3.2, p =0.12444 | NA | -1.54, p =1 |
| **Ex** | 1.48, p =1 | 2.6, p =0.84345 | 0.2, p =1 | -1.16, p =1 | 0.06, p =1 | -0.31, p =1 | 0.98, p =1 | 0.46, p =1 | -0.65, p =1 | NA |

***STable 1.*** Z and P_Bonf_ values for effects of age on directed connectivity on the edge-level.

|  | **VO** | **DMN** | **VM** | **VL** | **FPR** | **FPL** | **SM** | **Cer** | **Au** | **Ex** |
| --- | --- | --- | --- | --- | --- | --- | --- | --- | --- | --- |
| **VO** | NA | -3.87, p =0.00964 | 1.05, p =1 | 0.55, p =1 | -1.23, p =1 | -0.63, p =1 | 0.98, p =1 | 1.44, p =1 | 1.98, p =1 | -0.65, p =1 |
| **DMN** | -2.26, p =1 | NA | 2.18, p =1 | -0.9, p =1 | -1.36, p =1 | -1.45, p =1 | 0.12, p =1 | -0.16, p =1 | 0.76, p =1 | -1.15, p =1 |
| **VM** | 1.13, p =1 | -0.44, p =1 | NA | -1.23, p =1 | -1.3, p =1 | -1.18, p =1 | 0.15, p =1 | 0.62, p =1 | -0.58, p =1 | -1.17, p =1 |
| **VL** | -0.4, p =1 | -1.25, p =1 | -0.9, p =1 | NA | -1.37, p =1 | 0.05, p =1 | 0.4, p =1 | 0.3, p =1 | 1.46, p =1 | -1.08, p =1 |
| **FPR** | 0.07, p =1 | -1.8, p =1 | 1.14, p =1 | -0.48, p =1 | NA | -1.38, p =1 | 1.73, p =1 | 0.71, p =1 | 2.77, p =0.51102 | 1.51, p =1 |
| **FPL** | -0.39, p =1 | -1.84, p =1 | 0.06, p =1 | 0.22, p =1 | -1.88, p =1 | NA | 1.48, p =1 | -0.26, p =1 | 2.03, p =1 | -0.52, p =1 |
| **SM** | -0.81, p =1 | -2.64, p =0.7516 | -0.75, p =1 | -1.26, p =1 | -1.51, p =1 | 0.19, p =1 | NA | 0.57, p =1 | 1.56, p =1 | -0.58, p =1 |
| **Cer** | 0.07, p =1 | -1.9, p =1 | -0.65, p =1 | -0.04, p =1 | -2.12, p =1 | -0.15, p =1 | 1.57, p =1 | NA | 1.06, p =1 | -0.13, p =1 |
| **Au** | 0.23, p =1 | 0.2, p =1 | -0.27, p =1 | -0.74, p =1 | 0.31, p =1 | -1.05, p =1 | 2.26, p =1 | 0.33, p =1 | NA | -1.54, p =1 |
| **Ex** | 0.84, p =1 | -1.95, p =1 | 1.23, p =1 | -2.46, p =1 | -0.37, p =1 | -1.43, p =1 | 0.95, p =1 | 0.49, p =1 | 1, p =1 | NA |

***STable 2.*** Z and P_Bonf_ values for effects of FSIQ on directed connectivity on the edge-level.

|  | **VO** | **DMN** | **VM** | **VL** | **FPR** | **FPL** | **SM** | **Cer** | **Au** | **Ex** |
| --- | --- | --- | --- | --- | --- | --- | --- | --- | --- | --- |
| **VO** | NA | 3.71, p =0.01891 | 1.02, p =1 | 0.81, p =1 | 0.46, p =1 | 0.53, p =1 | 4.67, p =0.00027 | -0.62, p =1 | 0.46, p =1 | -0.81, p =1 |
| **DMN** | 3.03, p =0.22255 | NA | 1.35, p =1 | 3.28, p =0.09342 | -0.38, p =1 | 1.19, p =1 | 2.98, p =0.25994 | 0.16, p =1 | -0.84, p =1 | -1.47, p =1 |
| **VM** | 1.91, p =1 | 6.17, p =0 | NA | -0.4, p =1 | -2.8, p =0.45732 | -1.37, p =1 | -1.34, p =1 | 0.61, p =1 | -1.26, p =1 | -3.43, p =0.05405 |
| **VL** | -0.62, p =1 | 6.1, p =0 | -0.63, p =1 | NA | -0.87, p =1 | -0.75, p =1 | 0.33, p =1 | 1.93, p =1 | -0.49, p =1 | -1.39, p =1 |
| **FPR** | 1.43, p =1 | 5.42, p =1e-05 | -1.25, p =1 | 1.37, p =1 | NA | -0.54, p =1 | 2.71, p =0.60057 | 2.3, p =1 | 0.76, p =1 | -0.87, p =1 |
| **FPL** | 3.19, p =0.12874 | 4.39, p =0.001 | -0.61, p =1 | -1.03, p =1 | -1.65, p =1 | NA | 1.3, p =1 | 0.84, p =1 | -0.64, p =1 | 1.03, p =1 |
| **SM** | 4.03, p =0.00498 | 4.11, p =0.00355 | -2.3, p =1 | 0.29, p =1 | 0.94, p =1 | 0.81, p =1 | NA | 0.97, p =1 | -2.9, p =0.33494 | -3.85, p =0.01069 |
| **Cer** | -0.05, p =1 | 2.55, p =0.97566 | 2.1, p =1 | 2.25, p =1 | 1.59, p =1 | 2.73, p =0.56732 | 4.3, p =0.00155 | NA | -4.15, p =0.00301 | 1.62, p =1 |
| **Au** | 1.17, p =1 | 3.2, p =0.12457 | 1.01, p =1 | 0.67, p =1 | 1.3, p =1 | -0.1, p =1 | -0.14, p =1 | -3.31, p =0.08333 | NA | -2.17, p =1 |
| **Ex** | 0.34, p =1 | 4.83, p =0.00012 | -1.97, p =1 | -2.4, p =1 | -1.22, p =1 | 0.43, p =1 | 0.04, p =1 | 0.93, p =1 | -2.63, p =0.77463 | NA |

***STable 3.*** Z and P_Bonf_ values for effects of motion on directed connectivity on the edge-level.

|  | **VO** | **DMN** | **VM** | **VL** | **FPR** | **FPL** | **SM** | **Cer** | **Au** | **Ex** |
| --- | --- | --- | --- | --- | --- | --- | --- | --- | --- | --- |
| **VO** | NA | 0.33, p =1 | 0.8, p =1 | -2.93, p =0.3072 | -5.48, p =0 | -4.21, p =0.00228 | -0.95, p =1 | -1.15, p =1 | -3.62, p =0.02631 | -4.26, p =0.00182 |
| **DMN** | 0.96, p =1 | NA | -2.97, p =0.26686 | -1.74, p =1 | -5.01, p =5e-05 | -2.07, p =1 | 0.59, p =1 | 1.6, p =1 | -3.83, p =0.01168 | -4.67, p =0.00028 |
| **VM** | 1.41, p =1 | -0.67, p =1 | NA | -2.67, p =0.68175 | -7.92, p =0 | -5.36, p =1e-05 | -2.79, p =0.48012 | -1.46, p =1 | -1.96, p =1 | -5.8, p =0 |
| **VL** | -2.87, p =0.36989 | -0.01, p =1 | -2.3, p =1 | NA | -6.57, p =0 | -4.11, p =0.00349 | -2, p =1 | 0.34, p =1 | -2.4, p =1 | -3.96, p =0.00664 |
| **FPR** | -0.02, p =1 | 2.58, p =0.88952 | -1.68, p =1 | 0.53, p =1 | NA | -6.06, p =0 | -0.92, p =1 | -1.06, p =1 | -4.96, p =6e-05 | -3.67, p =0.02223 |
| **FPL** | 0.37, p =1 | 1.34, p =1 | -2.02, p =1 | -2.08, p =1 | -7.17, p =0 | NA | -4.3, p =0.00154 | -4.86, p =0.00011 | -2.62, p =0.78356 | -3.03, p =0.22125 |
| **SM** | -0.21, p =1 | 1.39, p =1 | 0.68, p =1 | 0.43, p =1 | -5.01, p =5e-05 | -5.95, p =0 | NA | -1.78, p =1 | -3.37, p =0.06733 | -6.21, p =0 |
| **Cer** | -2.24, p =1 | 0.24, p =1 | -2.16, p =1 | -3.91, p =0.00822 | -6.62, p =0 | -7.23, p =0 | -1.79, p =1 | NA | -4.22, p =0.0022 | -4.27, p =0.00173 |
| **Au** | -1.92, p =1 | -1.15, p =1 | 1.18, p =1 | -1.6, p =1 | -7.89, p =0 | -4.73, p =2e-04 | -2.74, p =0.56091 | -0.43, p =1 | NA | -7.04, p =0 |
| **Ex** | -0.96, p =1 | 1.08, p =1 | -1.87, p =1 | -3.63, p =0.02551 | -7.29, p =0 | -4.35, p =0.00121 | -3.8, p =0.01288 | -1.61, p =1 | -6.8, p =0 | NA |

***STable 4.*** Z and P_Bonf_ values for effects of tsnr on directed connectivity on the edge-level.

|  | **VO** | **DMN** | **VM** | **VL** | **FPR** | **FPL** | **SM** | **Cer** | **Au** | **Ex** |
| --- | --- | --- | --- | --- | --- | --- | --- | --- | --- | --- |
| **VO** | NA | -2.69, p =0.64267 | -0.99, p =1 | -2.04, p =1 | 1.65, p =1 | 1.44, p =1 | -2.12, p =1 | -1.16, p =1 | -3.37, p =0.06665 | 0.05, p =1 |
| **DMN** | -3.63, p =0.02527 | NA | 2.9, p =0.33881 | 0.01, p =1 | 2.14, p =1 | 1.96, p =1 | 1.33, p =1 | 2.96, p =0.27795 | 2.54, p =1 | 4.33, p =0.00136 |
| **VM** | -1.82, p =1 | 1.42, p =1 | NA | 3.98, p =0.00617 | 3.22, p =0.11644 | 1.36, p =1 | -2.85, p =0.38981 | 0.1, p =1 | -0.68, p =1 | 2.29, p =1 |
| **VL** | -0.82, p =1 | -0.05, p =1 | 4.67, p =0.00027 | NA | 4.44, p =0.00081 | 1.39, p =1 | -2.89, p =0.34239 | 1.89, p =1 | 0.93, p =1 | 1.52, p =1 |
| **FPR** | -1.64, p =1 | -1.05, p =1 | 2.77, p =0.50788 | 1.59, p =1 | NA | 3.22, p =0.11367 | -1.56, p =1 | 1.49, p =1 | 1.31, p =1 | 2.78, p =0.48746 |
| **FPL** | -0.96, p =1 | -0.11, p =1 | 0.34, p =1 | -1.45, p =1 | 3.98, p =0.00632 | NA | -0.06, p =1 | 4.25, p =0.00195 | -0.15, p =1 | 0.8, p =1 |
| **SM** | -1.3, p =1 | 1.83, p =1 | -0.89, p =1 | -2.5, p =1 | 2.07, p =1 | 2.13, p =1 | NA | 2.77, p =0.50572 | 3.14, p =0.15027 | 0.25, p =1 |
| **Cer** | -1.79, p =1 | 1.22, p =1 | 2.59, p =0.85432 | 3.22, p =0.1162 | 4.62, p =0.00035 | 5.06, p =4e-05 | 1.81, p =1 | NA | 1.36, p =1 | 6.04, p =0 |
| **Au** | -5.2, p =2e-05 | 1.62, p =1 | -0.62, p =1 | -1.47, p =1 | 3.1, p =0.17228 | 0.22, p =1 | -1.36, p =1 | 2.04, p =1 | NA | 3.17, p =0.13754 |
| **Ex** | -0.38, p =1 | 2.76, p =0.5153 | 1.99, p =1 | -0.42, p =1 | 4.82, p =0.00013 | 1.25, p =1 | -2.87, p =0.36434 | 5.14, p =2e-05 | 2.09, p =1 | NA |

***STable 5.*** Z and P_Bonf_ values for effects of scanner:Rutgers on directed connectivity on the edge-level.

|  | **VO** | **DMN** | **VM** | **VL** | **FPR** | **FPL** | **SM** | **Cer** | **Au** | **Ex** |
| --- | --- | --- | --- | --- | --- | --- | --- | --- | --- | --- |
| **VO** | NA | 4.15, p =0.00305 | -2.88, p =0.36194 | -1.29, p =1 | 1.38, p =1 | 0.11, p =1 | 3.91, p =0.00817 | 0.15, p =1 | -0.57, p =1 | 0.46, p =1 |
| **DMN** | 0.64, p =1 | NA | -0.95, p =1 | -0.51, p =1 | 3.66, p =0.02237 | 3.01, p =0.23701 | 3.81, p =0.0126 | 0.87, p =1 | 2.98, p =0.25684 | 1.89, p =1 |
| **VM** | -4.81, p =0.00013 | 0.12, p =1 | NA | 2.07, p =1 | 1.38, p =1 | 3.84, p =0.01099 | 3.92, p =0.00786 | -1.11, p =1 | 0.83, p =1 | -0.93, p =1 |
| **VL** | -1.4, p =1 | 2.03, p =1 | 1.07, p =1 | NA | 3.99, p =0.00594 | 1.84, p =1 | 1.74, p =1 | 3.3, p =0.08743 | -0.71, p =1 | 1.67, p =1 |
| **FPR** | -1.62, p =1 | 2.62, p =0.79917 | 0.16, p =1 | 0.73, p =1 | NA | 2.16, p =1 | 1.64, p =1 | 3.21, p =0.1209 | 0.93, p =1 | 0.91, p =1 |
| **FPL** | -1.71, p =1 | 3.68, p =0.0208 | 2.74, p =0.55024 | 0.26, p =1 | 2.78, p =0.48309 | NA | 2.39, p =1 | -2.09, p =1 | 1.67, p =1 | -0.91, p =1 |
| **SM** | 0.6, p =1 | 4.84, p =0.00012 | 1.92, p =1 | -1.48, p =1 | 2.64, p =0.74999 | 2.37, p =1 | NA | -0.42, p =1 | 1.6, p =1 | -1.2, p =1 |
| **Cer** | 0.38, p =1 | 4.04, p =0.00486 | 1.8, p =1 | 6.21, p =0 | 7.07, p =0 | 1.9, p =1 | 3.96, p =0.00673 | NA | 2.25, p =1 | -0.72, p =1 |
| **Au** | -4.16, p =0.00292 | 3.34, p =0.07596 | 0.6, p =1 | -1.57, p =1 | 2.31, p =1 | 1.07, p =1 | 1.63, p =1 | 0.18, p =1 | NA | 1.55, p =1 |
| **Ex** | -1.01, p =1 | 4.27, p =0.00174 | -0.68, p =1 | -0.09, p =1 | 2.74, p =0.54904 | 1.18, p =1 | 0.27, p =1 | -2.13, p =1 | 2.78, p =0.49592 | NA |

***STable 6.*** Z and P_Bonf_ values for effects of scanner:StatenIsland on directed connectivity on the edge-level.

**5. Correlation among measurements**

We looked into possible multicollinearity between covariates included in the covariate analysis. We found tSNR and motion to be highly correlated as would be expected but none of the other covariates were found to be highly correlated with each other (see figure S3).

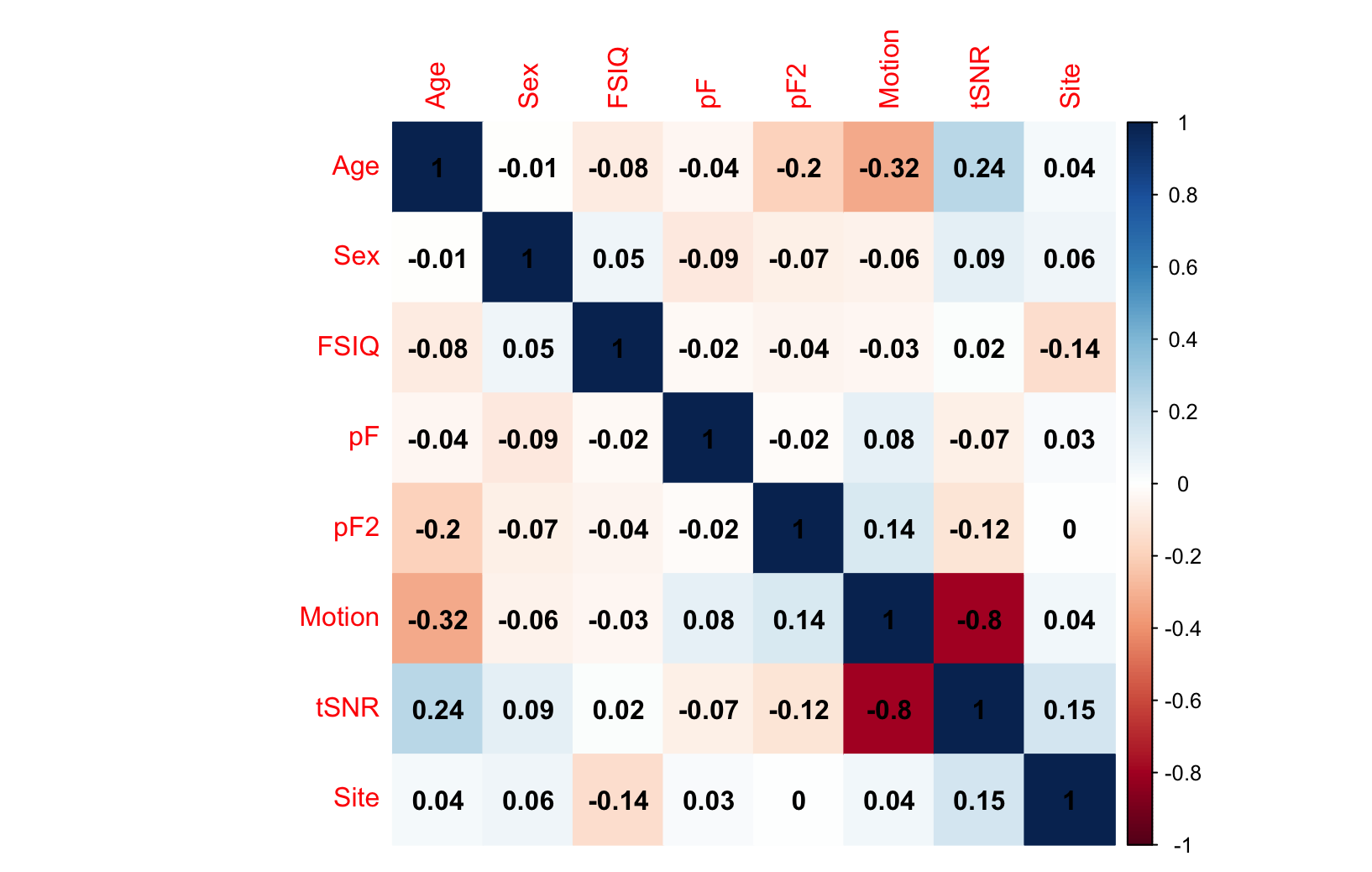

**Figure S5.** *Correlation between the covariates; age, sex, mental health, cognitive abilities, tSNR, site and motion included in the HBN model.*
